# Detection and upsurge of SARS-CoV-2 Omicron variant in Islamabad Pakistan

**DOI:** 10.1101/2022.01.13.22268997

**Authors:** Massab Umair, Aamer Ikram, Zaira Rehman, Syed Adnan Haider, Nazish Badar, Muhammad Ammar, Qasim Ali, Abdul Ahad, Rana Suleman, Ayesha Khalid, Amna Tariq, Zunera Jamal, Muhammad Salman

**Author notes:** **Corresponding author:** Massab Umair, Department of Virology, National Institute of Health, 45500, Park Road, Chak Shahzad, Islamabad, Pakistan.

## Abstract

The SARS-CoV-2 omicron variant was first detected in South Africa in November, 2021 and has rapidly spread to more than 90 countries. The emergence of Omicron variant demands for enhanced genomic surveillance to track the mutation profile and spread of virus. In the current study, we have sequenced 15 whole-genome sequences of SARS-CoV-2 Omicron variant from Islamabad region of Pakistan. Among the 15 isolates, 66% were from Islamabad whereas 33% of cases had international travel history of United Kingdom, Maldives, South Africa, and Oman. The detection of Omicron in local community and in travelers highlights the need for rigorous screening at national level and at entry points in order to contain the spread of variant.

## Introduction

Severe acute respiratory syndrome coronavirus 2 (SARS-CoV-2) was first identified in late December 2019, in Wuhan, China [1]. Subsequently, the virus has rapidly spread to several countries leading to an unprecedented pandemic that has disrupted normal life. As of January 9, 2022, 307 million cases and 5.49 million deaths have been caused by SARS-CoV-2 globally. During COVID-19 (coronavirus disease 2019) pandemic, SARS-CoV-2 has been evolving leading to emergence of different lineages and sub-lineages. Of note are variants of concern (VOCs) that are associated with increased transmissibility or increase in virulence or decreased effectiveness of public health and social measures or vaccines, therapeutics and diagnostics. Before November, 2021 four VOCs (alpha, beta, gamma and delta) were reported which contributed to new waves of infections in different countries. Genomic surveillance revealed the detection of a new variant of SARS-CoV-2 (B.1.1.529) from Botswana and South Africa in November 2021 which was immediately reported to the World Health Organization (WHO) on November 24, 2021. On November 26, 2021 this variant was declared as a variant of concern and named Omicron (B.1.1.529) by WHO based on the advice from Technical Advisory Group on Virus Evolution [2]. In South Africa, before the detection of Omicron the number of positive cases were averaging <1000 per day however, the rapid spread of Omicron resulted in increasing the number of cases with 23,283 cases reported on December 18, 2021. Similar increasing trend has been observed in United Kingdom and United States of America with 182, 890 and 574,507 number of positive cases reported respectively as of January 5, 2021 [3]. With COVID-19 cases soaring in USA and UK the variant has spread to more than 90 countries posing a major challenge in the control of the disease. In Pakistan, the first case of Omicron was detected on December 13, 2021 and since then COVID-19 cases have started to increase however, limited data is available on the genomic surveillance of Omicron from the country. The current study aims to explore the genetic epidemiology of Omicron in Islamabad region of Pakistan.

## Materials and Methods

### Sampling

The oropharyngeal swab specimens from COVID-19 suspected subjects (n=19,404) were collected during the month of December 2021 who visited the National Institute of Health as part of routine surveillance.

### RNA Extraction and real-time PCR

The MagMAX Viral/Pathogen Nucleic Acid Isolation kit (ThermoFisher Scientific, USA) and the KingFisher Flex instrument (ThermoFisher Scientific, USA) were used for RNA extraction. Clinical RT-PCR testing was performed using TaqPath^™^ COVID-19 RT-PCR kit (ThermoFisher Scientific, USA) that targets the three genes (ORF1ab, N, and S). In case of suspected Omicron cases, there was failure in amplification of S gene termed as Spike gene target failure (SGTF).

### Sample selection criteria for Whole genome sequencing

The SGTF samples with low cycle threshold values (≤30) were further confirmed for Omicron based on the positive amplification of K417N target and failure of E484K, and P681R mutation targets, using the SNPsig® SARS-CoV-2 (EscapePLEX) kit (PrimerDesign, UK). A subset of confirmed Omicron positive samples were selected for whole genome sequencing.

### cDNA synthesis, and Amplification

The cDNA synthesis and amplification was performed according to the ARTIC amplicon sequencing protocol (version 2) using SuperScript™ IV VILO™ Master Mix (Invitrogen, USA) and Q5® High-Fidelity 2X Master Mix (New England BioLabs, USA), with the ARTIC nCoV-2019 Panel V3 (Integrated DNA Technologies, Inc, USA) [4].

### Next Generation Sequencing

The paired-end sequencing library (2×150 bp) was prepared from the generated amplicons using the Illumina DNA Prep Kit (Illumina, Inc, USA) by following the standard protocol. The prepared libraries were pooled and subjected to sequencing on Illumina platform, iSeq using sequencing reagent, iSeq 100 i1 Reagent v2 (300-cycle) (Illumina, Inc, USA).

### Data Analysis

The Fastq files were processed for quality assessment using the FastQC tool (v0.11.9) [5]. Trimmomatic (v0.39) [6] was employed to eliminate artifacts and technical biases by removing adapter sequences and low-quality base calls (< 30). The filtered reads were aligned using the Burrows-Wheeler Aligner’s (BWA, v0.7.17) [7] and available reference genome (Wuhan-Hu-1, GISAID ID: EPI_ISL_402125). According to Centers for Disease Control and Prevention (CDC, USA) guidelines, variants were identified and consensus sequences for all genomes were generated [8]. Pangolin v3.1.17 and pangoLEARN model dated 06-12-2021 were used to classify the assembled genomes into PANGO lineages [9].

### Phylogenetic Analysis

The phylogenetic analysis of the study isolates was performed using the Ultafast Sample Placement of Existing Trees (UShER). It rapidly places the study samples onto existing phylogenetic tree of SARS-CoV-2 genomes reported on GISAID, NCBI, COG-UK, and CNBC using maximum parsimony method. The phylogenetic tree was constructed using the updated version on December 31, 2021. The resulting tree was visualized using Auspice.

## Results

During December 2022, a total of 19,404 samples were tested on real-time PCR for the presence of SARS-CoV-2 using the TaqPath^™^ COVID-19 kit (ThermoFisher Scientific, USA). Only 3% (n=679) of samples were positive for SARS-CoV-2.. Among the positive samples, 83% (n=566) showed amplification of spike gene, whereas 17% (n = 113) samples had the spike gene target failure (SGTF). On the basis of PCR-based VOCs genotyping assay (EscapePlex), spike gene positive (n=566) samples were characterized as delta whereas, SGTF samples (n=113) were found to be Omicron based on amplification of K417N and failure of E484K and P681R targets/markers. From these 113 Omicron samples, 15 samples (13%) were randomly selected for whole-genome sequencing (**Figure 1)**.

**Figure 1:**
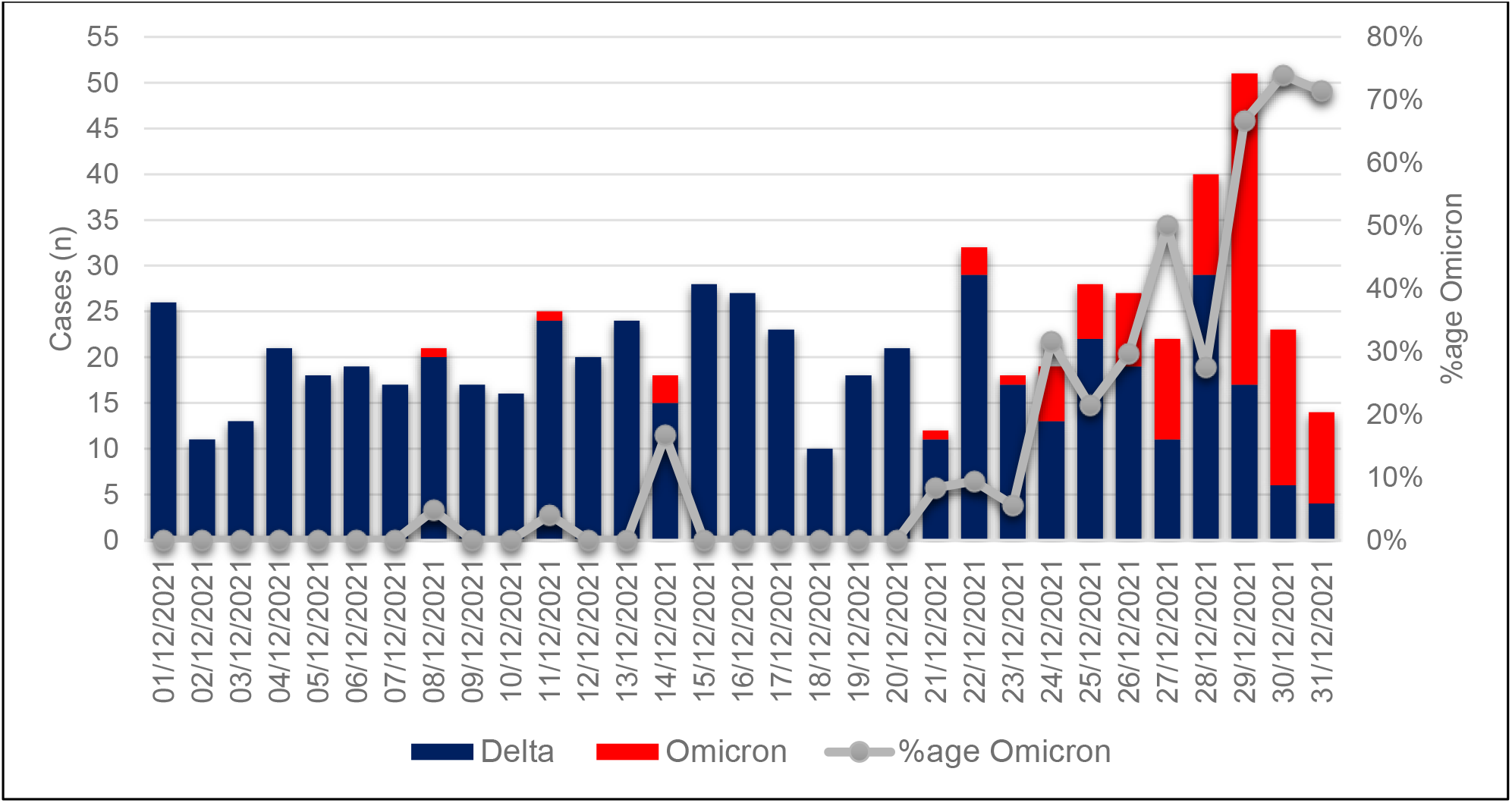
The data of genotyping COVID-19 positive cases during Decmber, 2022. The increasing trend in omicron positive cases was observed after December, 22, 2022.

The whole genome sequencing confirmed the presence of omicron variant (BA.1) in all the 15 isolates from Pakistan. The median age of patients was 36 years with age range of 20 to 67 years. The male to female ratio was 2:3 with 40% males and 60% females With reference to Omicron variant circulation within the city, 66% (n=10) of the Omicron infected samples were from the local resident of Islamabad while 33% (n=5) were identified from incoming passengers. The inbound passengers infected with omicron had travel history with two from UK (GISAID IDs: EPI_ISL_8240498 and EPI_ISL_8240500) and one each from Maldives (GISAID ID: EPI_ISL_8240512), Oman (GISAID ID: EPI_ISL_8240501) and South Africa (GISAID ID: EPI_ISL_8240513).

In all the 15 BA.1 isolates, the characteristic mutations of omicron variant was observed as follows: ORF1a:K856R, nuc:C3037T, nuc:T5386G, ORF1a:A2710T, ORF1a:T3255I, ORF1a:P3395H, ORF1a:I3758V, ORF1b:P314L, ORF1b:I1566V, S:A67V, S:T95I, S:G339D, S:S371L, S:S373P, S:K417N, S:N440K, S:G446S, S:S477N, S:T478K, S:E484A, S:Q493R, S:G496S, S:Q498R, S:N501Y, S:T547K, S:D614G, S:H655Y, S:N679K, S:P681H, S:N764K, S:D796Y, S:N856K, S:Q954H, S:N969K, E:T9I, M:D3G, M:Q19E, M:A63T, and N:RG203KR. Some rare spike mutations were also observed among the sequences as R346K in two of the isolates (GIASAID ID: EPI_ISL_8240504 and EPI_ISL_8240512 (travel history of Maldives)), A701V in four of the isolates (GISAID ID: EPI_ISL_8240500 (travel history of UK), EPI_ISL_8240506, EPI_ISL_8240507, EPI_ISL_8240508) and I1081V in only one isolate (GISAID ID: EPI_ISL_8240502). Phylogenetic analysis also revealed the introductions from England, Germany, Australia and USA. Despite the introductions, we also have community transmission cases (**Figure 2**).

**Figure 2:**
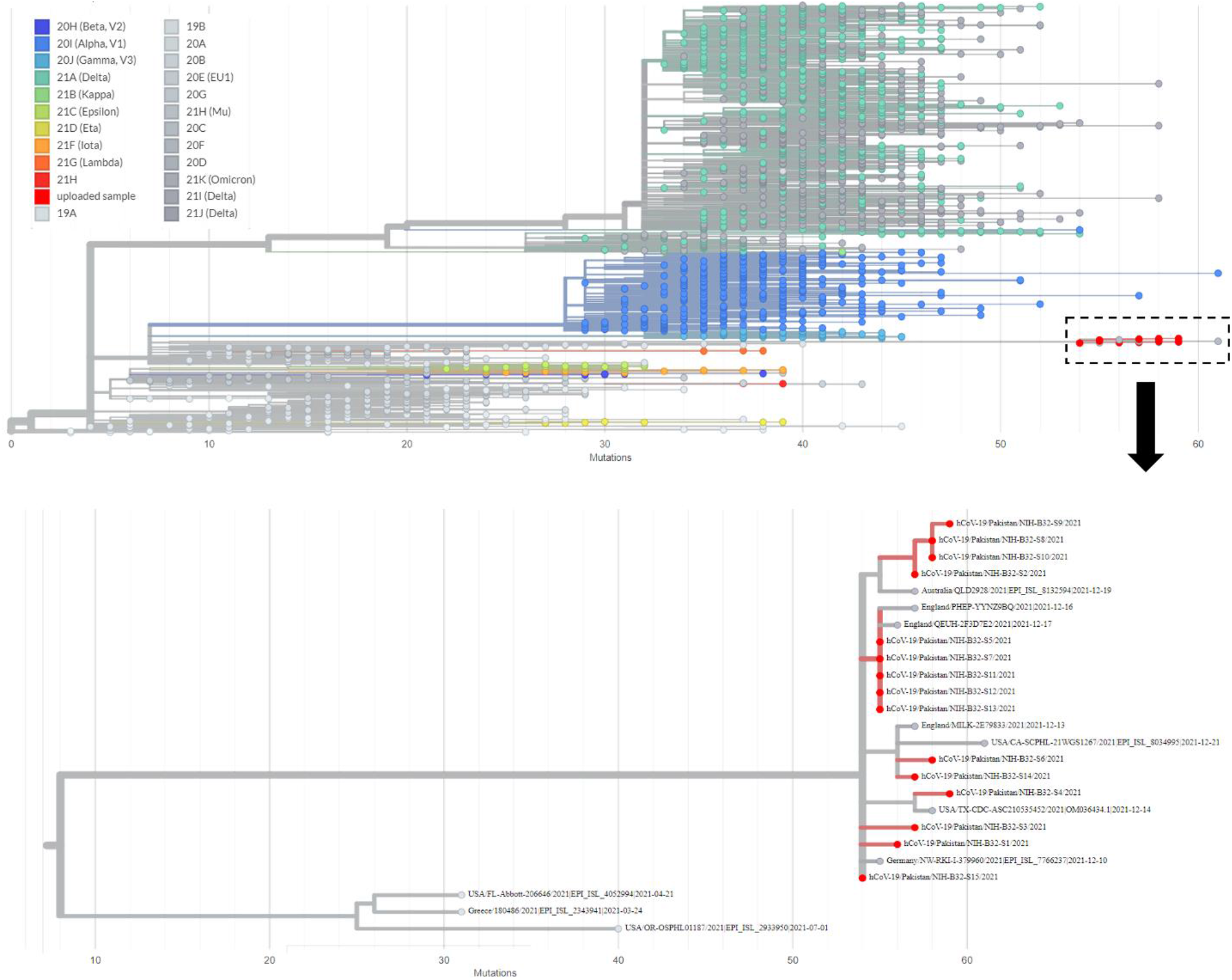
Phylogenetic tree of 15 Omicron isolates generated using USHER. The current study isolates as highlighted in red color.

## Discussion

The fourth wave of pandemic in Pakistan subsided in November, 2021 and till December 2021 COVID-19 cases remained below 700 per day. However, after the detection of first case of Omicron variant in Pakistan on December 13, 2021 the number of positive cases started increasing gradually. As of January 7, 2022 a total of 1,345 positive cases were reported in a single day [10]. The increasing trend in the number of positive cases is attributable to the spread of highly transmissible omicron variant in the country. As part of genomic surveillance the department of Virology, National Institute of Health identified 113 omicron positive cases in Islamabad. Further confirmation for the presence of omicron was made through whole-genome sequencing and 15 omicron whole genomes from SARS-CoV-2 positive patients were sequenced. There were 5 patients in current study that had a travel history to Maldives and United Kingdom. The phylogenetic analysis of Omicron from travelers showed introductions of the variant in Pakistan and highlights the need for screening of travelers at the points of entry for the timely isolation of such cases. This has been the case in most countries witnessing Omicron surge where the major share in Omicron cases is contributed by travelers from different countries. These travelers are the major source of importations from different countries and later results in community transmission.

The emergence of Omicron variant has raised alarms for a potential 5^th^ wave of COVID-19 in Pakistan. In light of the country’s sensitivity to a surge in omicron cases, it is critical to examine the genomic profile of the omicron circulating in Pakistan, as well as its mutational characteristics, to understand how the virus is evolving. Whether the mutations in the omicron variant can affect already vaccinated individuals as well as those who have not been vaccinated requires a continuous surveillance. Furthermore, implementation of travel restrictions, distancing measures, and promotion of mask wearing at all community places will be crucial which may aid in slowing down the COVID-19 disease transmission. Validation of these measures can be conducted with large-scale genomic surveillance from disease hotspots.

## Data Availability

All data produced in the present work are contained in the manuscript.

## Data Availability

All the sequences generated in the current study are submitted to the GISAID that are available at “https://www.gisaid.org/login/” under the accession numbers: EPI_ISL_8240498, EPI_ISL_8240500-EPI_ISL_8240513.

